# AIBx, artificial intelligence model to risk stratify thyroid nodules

**DOI:** 10.1101/2021.02.13.21251688

**Authors:** Johnson Thomas, Tracy Haertling

**Affiliations:** Department of Endocrinology, Mercy Hospital, Springfield, Missouri; Mercy Research Springfield, Missouri

**Keywords:** Artificial intelligence, Image similarity, thyroid nodule, thyroid cancer

## Abstract

**Background:** Current classification systems for thyroid nodules are very subjective. Artificial intelligence (AI) algorithms have been used to decrease subjectivity in medical image interpretation. 1 out of 2 women over the age of 50 may have a thyroid nodule and at present the only way to exclude malignancy is through invasive procedures. Hence, there exists a need for noninvasive objective classification of thyroid nodules. Some cancers have benign appearance on ultrasonogram. Hence, we decided to create an image similarity algorithm rather than image classification algorithm.

**Methods:** Ultrasound images of thyroid nodules from patients who underwent either biopsy or thyroid surgery from February of 2012 through February of 2017 in our institution were used to create AI models. Nodules were excluded if there was no definitive diagnosis of benignity or malignancy. 482 nodules met the inclusion criteria and all available images from these nodules were used to create the AI models. Later, these AI models were used to test 103 thyroid nodules which underwent biopsy or surgery from March of 2017 through July of 2018.

**Results:** Negative predictive value of the image similarity model was 93.2%. Sensitivity, specificity, positive predictive value and accuracy of the model was 87.8%, 78.5%, 65.9% and 81.5% respectively.

**Conclusion:** When compared to published results of ACR TIRADS and ATA classification system, our image similarity model had comparable negative predictive value with better sensitivity specificity and positive predictive value. By using image similarity AI models, we can eliminate subjectivity and decrease the number of unnecessary biopsies. Using image similarity AI model, we were able to create an explainable AI model which increases physician’s confidence in the predictions.

## Introduction

Ubiquitous use of imaging modalities for evaluation of various medical conditions leads to the discovery of incidentalomas. Being present in more than half of women over age 50, thyroid nodules are common incidentalomas. Analysis of Medicare data (1) showed that thyroid ultrasound as the initial imaging modality in the cohort has risen by 20.9% year over year.

Current classification systems for thyroid nodules are labor intensive and are subjective (2). The most common systems used to classify thyroid nodules are TIRADS and American Thyroid Association (ATA) classification system (3, 4). These systems are fraught with problems. Varying results can be seen when different classification systems are used to assess the same thyroid nodule. The ability to make a useful distinction especially by less experienced users is limited by the inherent subjectivity and the inter and intra reader variability of these visual classification systems. Using the above systems, Follicular carcinomas, Hurthle cell cancer and follicular variant of papillary thyroid cancer may end up being classified as benign (5). Not all nodules can be classified using all available systems. These classification systems also lack specificity and have low positive predictive value (6). This results in unnecessary biopsies. Millions of thyroid biopsies are done every year all over the world. It was estimated that in 2015, more than 600,000 fine needle aspirations (FNA) were done in the United States alone (7). Evaluation of the increasing number of benign thyroid incidentalomas is increasing the burden on the healthcare system.

Even when FNA of the thyroid is performed it does not always yield a definitive result. A final diagnosis cannot be made in one out of seven nodules with FNA (8). Molecular markers were developed to avoid surgery for benign nodules with indeterminate cytology. The positive predictive value for these molecular tests varies between 20 to 50% (8, 9). Many times, a repeat biopsy may be required to do molecular markers. All of this adds to healthcare expense without improving morbidity or mortality. Another possible outcome of FNA is a non diagnostic cytology report. Management options for this scenario includes repeat biopsy, surgery or watchful waiting. False negative results can occur after a biopsy and it is estimated that less than 5% of cytologically benign nodules are proved to be malignant after surgery. However, only about 10% of cytologically benign nodules undergo surgery, hence accurate estimates of false negative results may not be possible (7, 10, 11). Therefore, at present, we do not have a reliable non subjective method for avoiding invasive procedures for benign thyroid nodules.

Similar problems exist in other medical domains and artificial intelligence (AI) algorithms have provided solutions. There are FDA cleared AI software to diagnose diabetic retinopathy, stroke, and breast lesions (12–15). AI algorithms have been used to classify thyroid nodules objectively (16–19). Given a thyroid ultrasound image, these algorithms can predict whether a thyroid nodule is benign or malignant. However, predictions from these algorithms are not generally explainable hence they are called black box algorithms. In clinical practice, explainable or interpretable deep learning models are needed to gain the trust of physicians (20). Deep learning algorithms may make predictions based on non-medically relevant information present in the images. Winkler et al. demonstrated that having gentian violet surgical skin markings in dermoscopic images increased the chance of melanoma prediction by an algorithm (21). Images obtained from different imaging machines will have different features. This can act as a confounding factor while creating deep learning algorithms. For example, if pneumonia is more common in x-rays obtained in the Emergency Room (ER) when compared to surgical ward, then there is an increased possibility of false positives on X-rays performed in ERs. Algorithms created in one institution may underperform when used in another institution. Algorithms may overlook the actual pathology and instead may rely on other clinically non relevant features in the image like placement of a metallic token to mark laterality.(22) Because of these shortcomings we decided to create an image similarity AI model instead of a classification model. AI image classification algorithms for thyroid nodules gives a single output, benign or malignant without any supporting evidence regarding how it reached that conclusion. On the other hand, an image similarity algorithm will output similar images to the test image with corresponding diagnosis.

In this article, we describe the creation of an image similarity deep learning algorithm for thyroid nodule risk stratification.

## Materials and methods

The research study was approved by the Mercy Institutional Review Board. Image database

Ultrasound images of thyroid nodules from patients who underwent either biopsy or thyroid surgery from February of 2012 through February of 2017 at Mercy Endocrinology Clinic or Mercy Hospital in Springfield, Missouri were initially collected for the study. Cytology and histopathology examinations were done at the Mercy hospital by one group of pathologists. Cytopathology was reported using the Bethesda system (23).

Nodules were excluded if there was no definitive diagnosis of benignity or malignancy or if there were no good quality thyroid ultrasound images. 482 nodules met the inclusion criteria. The area of interest was cropped from the ultrasound images along with some neighboring tissue. Both sagittal and transverse view images were used. This image set served as the training database. The testing imaging dataset was created in a similar fashion from the retrospective collection of ultrasound images of patients with thyroid nodules who underwent biopsy or surgery between March 2017 through July of 2018. There were 103 thyroid nodules in the testing dataset.

### Convolutional Neural Network model

A 34 layered Convolutional Neural Network (CNN) -ResNet 34 was trained on thyroid ultrasound images of 482 thyroid nodules using transfer learning techniques (24). All images were resized to 224 × 224 pixels before being fed into the CNN. Image embeddings for these ultrasound images were obtained by taking the output before the final fully connected layer and stored in a database. Embeddings are N dimensional vectors representing one unique image. When a query image is received, it is first converted to image embeddings using the CNN. Embeddings from the input image are used to find embeddings that are similar to it from our training image database using a nearest neighbor algorithm. Finally, N number of nearest neighbors will be displayed as the output along with the label of the image. Fig. 1 depicts the schema of image similarity algorithm.

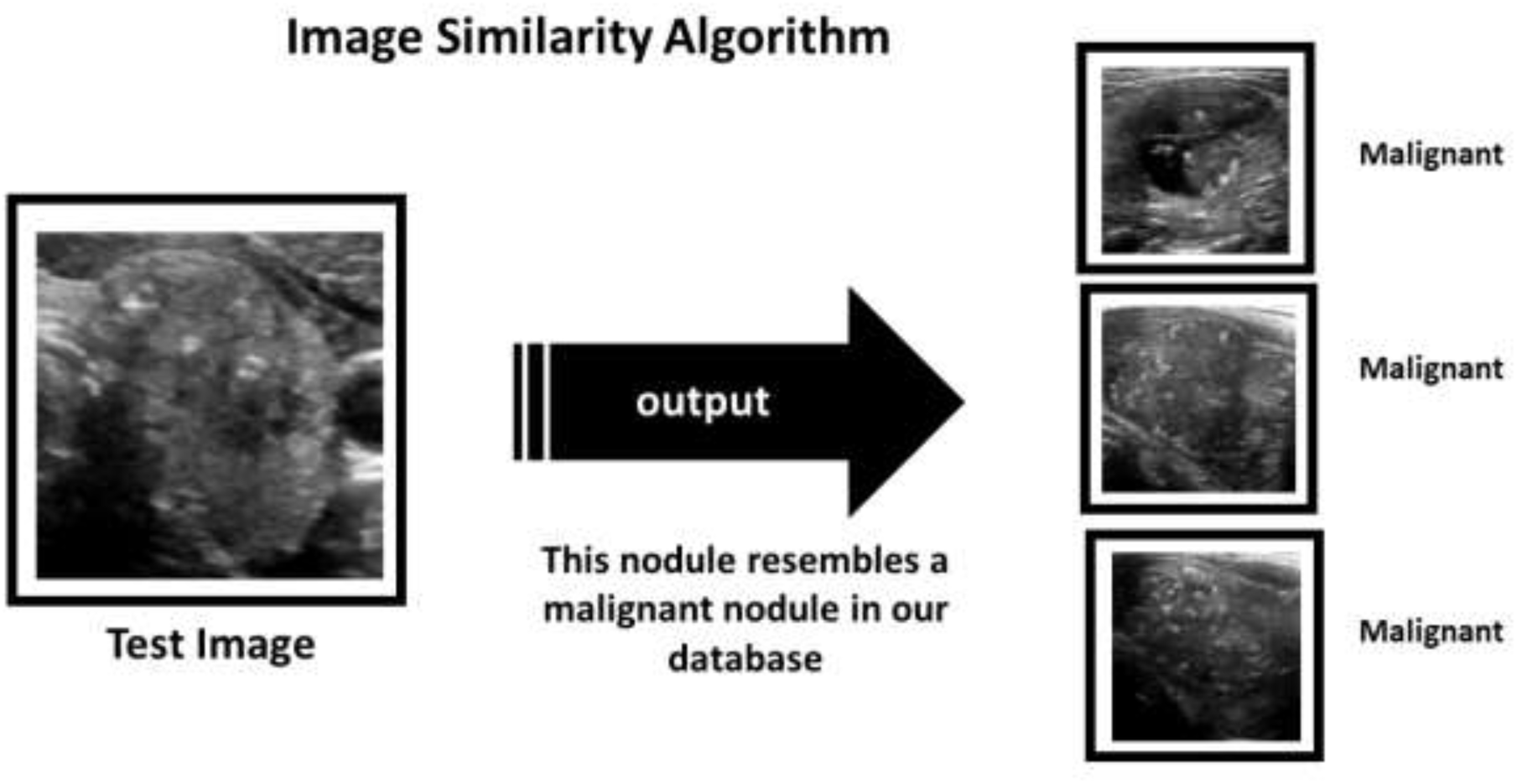

### Scoring the algorithms

In phase 1 image classification algorithms were used. The ResNet 34 model trained on 482 nodule images was used to classify test images. Algorithm returned a prediction for the test image as either benign or malignant. In phase 2, each of the test images were fed through the image similarity algorithm, AIBx. Image embeddings were created and the first nearest neighbor / similar image from our training dataset was identified. If the nearest neighbor for a benign test image is a benign nodule from the training database, it was considered a true negative. If the algorithm outputs a malignant nodule as the similar image for a malignant nodule in the test set it was considered a true positive. Opposite was true for false negative and false positives respectively.

### Statistical methods

A confusion matrix was created from the true positives, true negatives, false positive and false negatives in both phases. Python programming language was used to calculate accuracy, sensitivity, specificity, positive predictive value, and negative predictive value.

## Results

The training dataset consisted of 2025 images from 482 nodules. These included images with and without square aspect ratio. Testing set had 103 images from 103 nodules. Training and testing set had 66 and 33 malignant nodules respectively. Images used in the study came from ultrasound machines manufactured by GE, Siemens, Philips and Sonosite. Of the training set, 6% were subcentimeter nodules. All of the testing nodules had at least one dimension greater than 1 cm.

### Phase 1

Image classification using ResNet 34 model resulted in an accuracy of 77.7%. Sensitivity, specificity, positive predictive value, and negative predictive values were 84.9%, 74.3%, 60.9% and 91.2% respectively. The average time for prediction was 30 milliseconds per image.

### Phase 2

When the image similarity model was used to classify test Images, accuracy was 81.5%. The sensitivity, specificity, positive predictive value and negative predictive value of the model was 87.8%, 78.5%, 65.9% and 93.2% respectively. Average time for prediction was 900 milliseconds.

In using the image classification algorithm generated in phase 1, 55.3% of the nodules were determined to be benign. When the image similarity algorithm was used, 57.3% of the nodules were determined to be benign. Hence, using image similarity algorithm will avoid more biopsies. When results of Phase 1 and Phase 2 were compared (Table 1), image similarity algorithm turned out to be superior.

## Discussion

One of the main challenges in the management of thyroid nodules is risk stratification. With more than 50% of women over the age of 50 affected by thyroid nodules and millions of biopsies done every year resulting in less than 5 to 10% cancer diagnosis, a reliable noninvasive test is needed. Experienced physicians generally evaluates an ultrasound image and arrives at a decision regarding biopsy based on their previous experience and heuristics. Most have a mental picture of how a malignant thyroid nodule should appear. We tried to emulate this by creating an image similarity CNN model. While the repertoire of representative images stored in a physician’s mind is limited by his experience and memory, AI models can store unlimited images and query it millions of time.

Multiple artificial intelligence models have been developed for thyroid nodule classification, but none of them are widely used (17–19). A recent study by Buda et al suggested that machine learning algorithms could match the performance of radiologists in classifying thyroid nodules (16). When tested on 99 nodules, their model achieved a sensitivity of 87% and specificity was 52%. This was comparable to the performance of three ACR-TIRADS committee members and nine other radiologists (16). Most image classification algorithms used in the risk stratification of thyroid nodules are black boxes. Therefore, we cannot readily explain why an algorithm yielded the wrong classification of a thyroid nodule. Heatmaps have been used to explain outputs of image classification. But this approach does not help with thyroid nodule classification. Fig. 2A depicts a benign cystic nodule. Image classification algorithm correctly classified it as a benign nodule. The corresponding heatmap in Fig. 2A shows that the algorithm is focusing on the cystic area and the posterior enhancement to arrive at the diagnosis. Fig. 2B depicts a malignant thyroid nodule. This was classified as a benign nodule by image classification algorithm. But the heatmaps does not help us to understand the rationale behind this prediction.

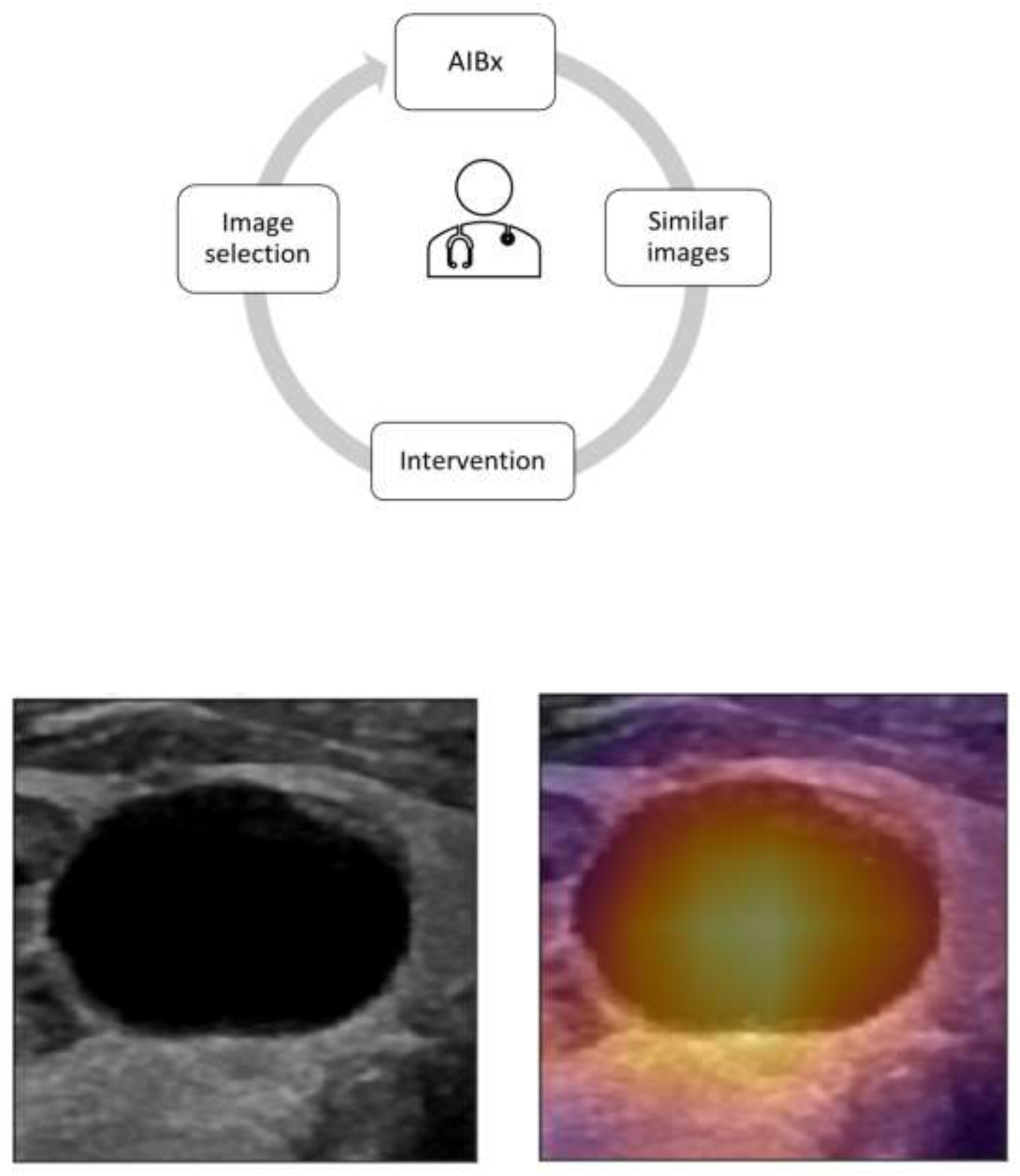

Explainable AI models will increase the trust of physicians and will foster adoption of these systems into clinical practice (20). AI healthcare team from Google created an image similarity model (SMILY) to help diagnose histopathology images (25). Given a histopathology image, SMILY can output images with similar histological features. They suggested that this approach could also inform us about the outcomes of patients with similar pathology.

To our knowledge, there is no published study on the use of image similarity models for the classification of thyroid. Unlike other AI algorithms, our image similarity model, AIBx uses physician in the loop (PIL), Fig. 3. During each stage of AIBx, physicians have an active role. Operating physicians will select the image to be fed into AIBx. AIBx will output N number of similar images as requested by the user along with a classification. Physicians can verify the diagnosis by reviewing the similar images and then accept or reject the classification provided by AIBx. This could also be used to retrieve all available information for a nodule including diagnosis, molecular markers, treatment received and recurrence status.

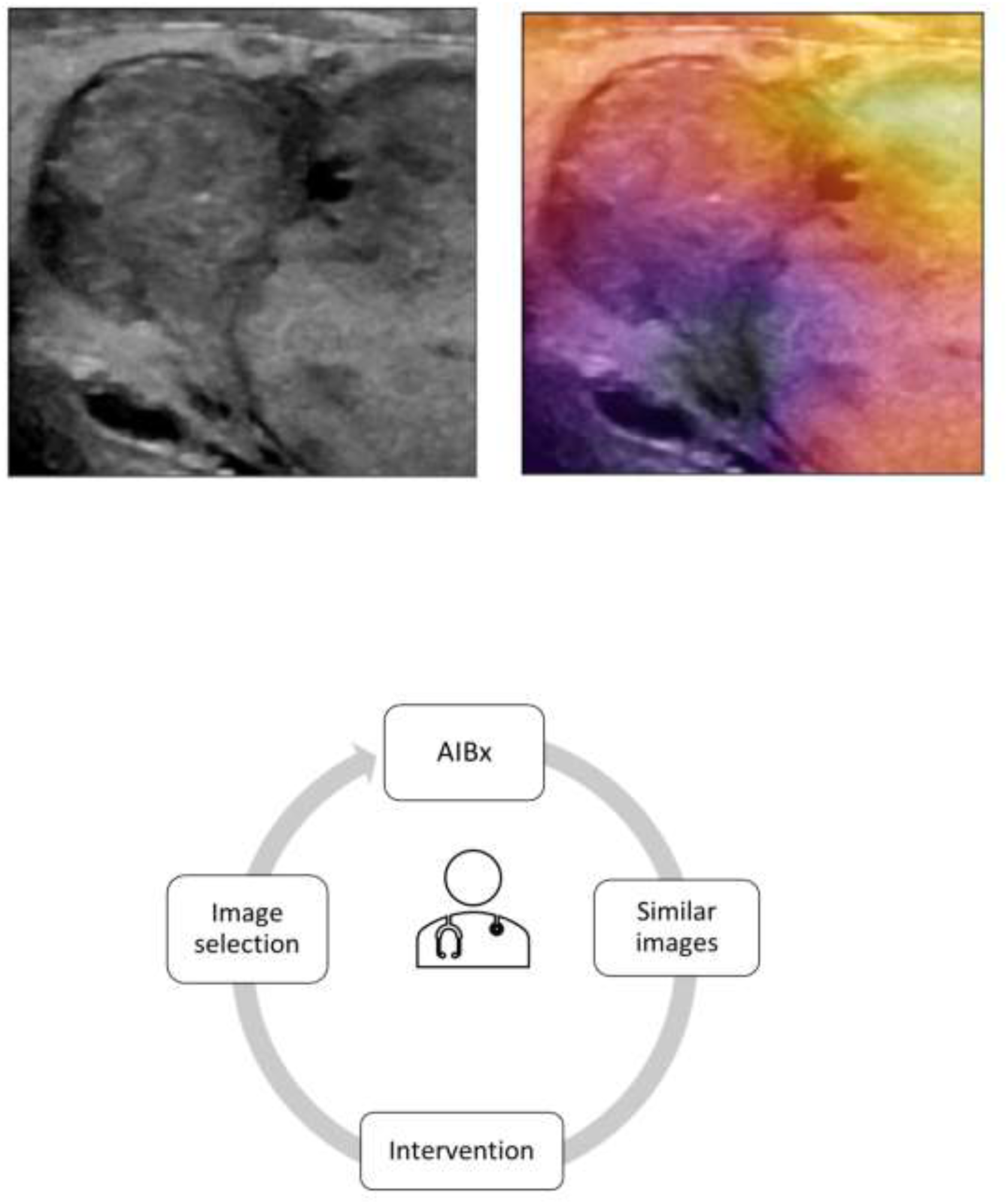

### Advantages

If an AI classification algorithm misclassified an image, adding this image back to the database and retraining the AI model may not result in the correct classification. Hence, retraining the classification model again with incorrectly classified images may not always result in better prediction of the misclassified images. However, a properly trained image similarity model has a high chance of reclassifying the image correctly if embeddings of the misclassified image are added to the database. Therefore, an image similarity model does not have to be retrained to increase accuracy. Models trained on images from one ultrasound machine may not generalize well to images classified from another ultrasound machine. AIBx used images from ultrasound manufacturers frequently used in clinical settings, including GE, Philips, Siemens and Sonosite. AIBx can easily be incorporated into current physician workflow. Any ultrasound machine with an image output port can wirelessly transmit the image of interest to a nearby mobile computing device where it can be classified using the algorithm. If this method is used, data never leaves the healthcare facility. AIBx can also be deployed as a local or remote website. Furthermore, the system could also be used as a teaching tool for residents and fellows.

### Potential disadvantages

Image similarity algorithms can consume more computing resources and time when compared to classification algorithms. But the difference was under a second for AIBx when compared to phase 1, image classification algorithm. The test dataset only had 103 images. It is possible that a larger test set may yield different results. Images acquired from an ultrasound machine other than the machines used in the study may not produce the correct response. However, this could be verified by the physician comparing similar images generated by AIBx to the test image. During testing, AIBx retrieved images from ultrasound machines other than the one from the test image, partly alleviating these concerns. Most thyroid nodules evaluated by FNA or surgery may have worrisome features and nodules that were not biopsied and/or surgically removed may have a benign appearance. As such, there could be underlying selection bias in our database.

A study by Grani et al. applied classification systems, ACR-TIRADS, ATA, AACE, EU-TIRADS and K-TIRADS to 502 nodules and reported that 11 malignant nodules would have been classified as not requiring biopsy by at least one of these systems (4, 6, 26–29). The ATA system could not classify some of the nodules (6). This study shows the variability in classification systems even with experienced physicians. According to this study, ACR-TIRADS performed better and recommended the lowest number of biopsies. The PPV and NPV for ACR-TIRADS was 12.8% and 97.8% respectively. In another study by Ahmadi et al. NPV for both ATA and ACR-TIRADS was 90% (30). All of these classification systems are subjective and will yield different results when applied by different practitioners. Using AIBx will eliminate subjectivity without significant compromise in negative predictive value. Thyroid cancer has a 98.2% 5-year survival rate and low morbidity (31). Combined with the practice of active surveillance of thyroid cancer by many centers, a system with greater than 90% negative predictive value will be helpful in avoiding unnecessary biopsies without increasing morbidity or mortality (32).

## Conclusion

Millions of thyroid biopsies are done every year based on very subjective criteria to find thyroid cancer in a very small percentage of population with an invasive technique which may not be diagnostic 1 out of 7 times. Here we described an image similarity algorithm based on deep learning for thyroid nodule risk stratification. When compared to published results of ACR TIRADS and ATA classification system, AIBx, the image similarity model had comparable negative predictive value with better sensitivity specificity and positive predictive value. By using image similarity AI models, we can eliminate subjectivity and decrease the number of unnecessary biopsies. This algorithm may also aid in the management of indeterminate and non-diagnostic thyroid nodules. Using image similarity AI model, we were able to create an explainable AI model which encourages physician’s confidence in model predictions.

## Data Availability

Data used for statistical analysis is available upon request.

## Acknowledgments

We thank Mercy research for facilitating the study.

